# Stress internalization associated with cognitive decline among older U.S. Chinese

**DOI:** 10.1101/2024.02.21.24301736

**Authors:** Michelle H Chen, Yiming Ma, Charu Verma, Stephanie Bergren, XinQi Dong, William T Hu

## Abstract

**INTRODUCTION:** Behavioral and sociocultural factors in minority populations in the U.S. are commonly examined as independent contributors or buffers to health disparities in cognitive decline, but can demonstrate significant inter-relatedness due to broader contextual factors (e.g., acculturation). Analyzing influence of correlated behavioral and sociocultural traits can better identify targets for future intervention. The current study aimed to account for interdependent risk and resilience factors associated with cognitive decline in non-demented older U.S.-dwelling Chinese adults.

**METHODS:** The sample consisted of 1,528 older U.S. Chinese (60 and older) with normal cognition and function at baseline who attended three waves of the Population Study of ChINese Elderly (PINE). We used principal component analysis to generate memory and executive functioning outcomes; factor analysis to reduce behavioral and sociocultural variables into latent constructs; and linear mixed-effects models to evaluate the impact of these constructs, as well as of demographic and medical factors (e.g., cardiovascular disease), on longitudinal rates of cognitive decline.

**RESULTS:** Factor analysis identified three main behavioral/sociocultural constructs: stress internalization, neighborhood/community cohesion, and external social support. Among these, only stress internalization – consisting of greater perceived stress, greater hopelessness, and lower conscientiousness – was associated with longitudinal decline in memory, while none with decline in executive functioning. Neither acculturation nor social engagement was related to decline in memory or executive function, even though participants with greater acculturation or social engagement had better baseline cognitive performance.

**DISCUSSION:** Using a psychometrically and statistically robust model, we found that only the factor underlying stress processing, hopelessness, and conscientiousness was associated with rates of memory decline in this older non-demented U.S. Chinese cohort. These maladaptive traits have been linked to the Asian model minority stereotype but all the same potentially modifiable. Future studies examining disparate health outcomes must account for inter-relatedness among behavioral and sociocultural factors to identify root causes.

## 1. Introduction

The number of older Asian Americans is expected to grow 93% by 2040.^1^ Among ethnic Asian groups, U.S. Chinese (individuals of Chinese descent living in the U.S.) represent the largest subgroup and constitute 24% of all Asian Americans.^2^ Older U.S. Chinese – similar to other older racial/ethnic minority populations – are consistently underrepresented in memory and aging studies,^3^ yet available studies suggest older U.S. Chinese face increased risk of delayed diagnosis and treatment of dementia.^4^ This can result from reduced knowledge of dementia as an illness rather than an inevitable aspect of aging,^5,6^ cultural stigma associated dementia and other mental illnesses,^7,8^ and stronger dyadic or family interdependence which can mask functional decline.^9,10^ The largely immigrant (69%)^11^ U.S. Chinese population may also have health-related knowledge which has remained stagnant from their time of immigration, and their access to lay^12^ or professional^13,14^ sources of information could be made worse by 67% of older U.S. Chinese speaking English very poorly or not at all.^11^ Even though common health prevention approaches may provide cognitive benefits across racial/ethnic groups, there is a paucity of direct evidence linking factors targeted by these approaches to rates of cognitive decline among U.S. Chinese.

The Chicago-based **<u>P</u>**opulation Study of Ch**<u>IN</u>**ese **<u>E</u>**lderly (PINE) is the largest community-based cohort study on older U.S. Chinese. Unlike earlier studies on cognition among Asian Americans such as the Honolulu-Asia Aging Study (HAAS)^15^ and the Multi-Ethnic Study of Atherosclerosis (MESA),^16^ PINE participants represented a smaller ethnic group (Chinese Americans in the PINE vs. Japanese Americans in the HAAS who represent 25-45% of the Hawaii population throughout the 20^th^ Century^17^), were more likely to live in an ethnic enclave (1/3 of Chicago’s Chinese population live in or near its Chinatown), and had lower socioeconomic status (23% of PINE participants vs. 64% of Chinese MESA participants had at least some college education). These distinctions make PINE a unique resource to examine the impact of sociobehavioral factors on cognition.

Prior PINE studies – often examining a select number of risk or resilience factor and a single mean cognitive score – correlated multiple measures of healthier living (e.g., social engagement, cognitive leisure activities) with better cognition.^18–22^ However, repeatedly examining only subsets of factors increases cohort-level Type I errors. Multiple sociobehavioral variables in immigrant populations can also be jointly influenced by internal (e.g., acculturation) or external (e.g., access to transportation) factors. This interdependence between variables is not new in biomedical research, but is better accounted in genomic, proteomic, and other big data approaches through haplotypes (e.g., genes inherited in a related fashion) and eigenvectors (e.g., proteins regulated in a related fashion). For outcomes, prior PINE studies operationalized cognitive decline as a one-way crossing beyond a threshold (Z-score of −1.5) of a participant’s average performance in five short tests relative to the entire cohort.^23^ The potential inclusion of people with mild baseline impairment during derivation of this threshold risks underestimating the proportion of people experiencing meaningful cognitive decline, and the threshold approach overlooks common fluctuations in cognitive performance in population studies. Together, conceptual and methodological shortcomings can conflate statistical anomalies with meaningful associations which in turn worsen health disparities among older U.S. Chinese.

To best prioritize actionable risk and resilience factors in age-associated cognitive decline among older U.S. Chinese, the current paper will address limitations of prior PINE studies by identifying participants with normal cognition at baseline, assessing longitudinal cognitive trajectories, and accounting for interdependence among sociobehavioral risk factors.

## 2. Methods

### 2.1. Ethics in Human Research

This study was approved by the Institutional Review Boards of Rush University Medical Center and Rutgers University. The study was performed in accordance with ethical standards laid down in the 1964 Declaration of Helsinki and its later amendments, and signed informed consents were obtained from all participants.

### 2.2. Diversity, Equity, and Inclusion

The PINE investigators worked intensively with a community advisory board (CAB), consisting of diverse community stakeholders across 20 organizations in the Chicago metropolitan area, in designing the study and subject recruitment.^24^ The measures were translated and back-translated by an experienced bilingual and bicultural research team into both simplified and traditional Chinese characters.^23^ Since the study focused on older U.S. Chinese, participants were all of Chinese descent, although country of origin can vary across multiple countries. To ensure diversity of Chinese subcultures, study interviews were offered in both Mandarin and Cantonese, and there was a concerted effort to recruit approximately equal number of men and women as well as individuals with disabilities.

### 2.3. Participants

Study sample was drawn from waves 1-3 of PINE. Study design and participant characteristics were previously detailed.^24,25^ Briefly, PINE is a community-based epidemiological study of older adults of Chinese descent (aged 60 and older) living in the greater Chicago, Illinois area (midwestern U.S.). In-person interviews were conducted in 2011-2013, 2013-2015, and 2015-2017 for the three waves. 1,528 subjects who were non-demented at baseline and attended all three waves were included in this study (see Figure 1 for participant selection).

**Figure 1.**
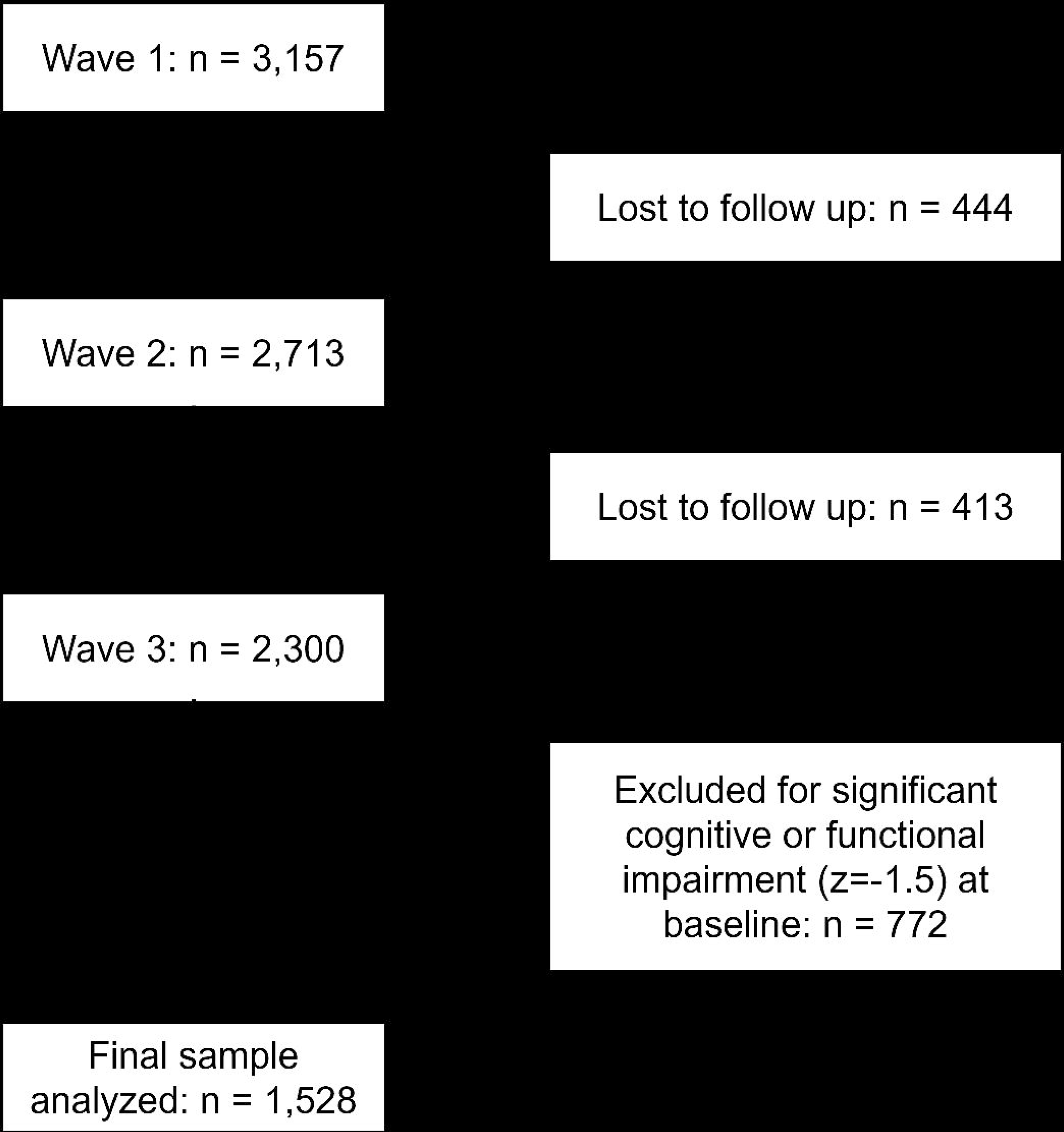
CONSORT diagram.

### 2.4. Measures

See Supplementary Table 1 for comprehensive information about all study measures.

#### 2.4.1. Cognitive functioning

Five cognitive measures were collected in PINE: Global cognitive functioning by the Chinese version of the Mini-Mental State Examination (C-MMSE),^26^ episodic memory by the East Boston Memory Test (EBMT immediate and delayed recall),^27^ working memory by the Wechsler Memory Scale-Revised (WMS-R)^28^ Digit Span Backward subtest, and processing speed by the oral version of the Symbol Digit Modalities Test (SDMT).^29^

#### 2.4.2. Potential predictors of cognitive functioning

Demographic variables included age at wave 1, sex, years of formal education, marital status, and annual income. Acculturation was measured by the PINE Acculturation Scale.^30^ Current and past health were assessed by self-reported diagnosis of a list of health conditions (Supplementary Table 1), along with a semi-quantitative sum of four cardiovascular risks (heart disease, stroke, high blood pressure, and diabetes). Symptoms related to 15 organ systems and smoking history were also assessed. Depression,^31,32^ anxiety,^33^ conscientiousness/neuroticism,^34^ perceived stress,^35^ hopelessness,^36^ neighborhood cohesion,^37^ sense of community,^38^ perceived social support,^39^ and social activity engagement^40^ were assessed using standardized self-report inventories (Supplementary Table 1).

### 2.5. Statistical Analyses

See Supplementary Table 1 for details regarding data transformations. Data analyses were conducted using IBM SPSS version 28 and plots were generated using R version 4.3.1.

#### 2.5.1. Cognitive outcomes

To address the potential inclusion of participants with baseline cognitive impairment in the derivation of Z-scores, we established a normative subgroup within PINE. We first identified participants with C-MMSE>28 at wave 3 which excluded incident dementia while allowing for practice effects during early waves. To additionally account for the possibility that some cognitively impaired individuals had high C-MMSE, we reduced heterogeneity in this normal cohort by removing individuals who had Lawton-Brody Instrumental Activities of Daily Living (IADL)<7 (equivalent to Z-score of −1.5 as there was no difference between men and women in this cohort) during any wave. The resultant subgroup’s performance at wave 1 (n=724) formed the normative curve for each cognitive measure. Raw performance also underwent linear regression analysis to adjust for age, sex, and education before Z-transformation to produce adjusted Z-scores. All participants’ Z-scores across the five measures were then analyzed using principal component analysis (PCA; Varimax rotation), arriving at two principal components (PC) whose scores were used as cognitive outcomes.

#### 2.5.2. Predictors

To test the non-independent nature of sociobehavioral risk/resilience factors, we conducted exploratory factor analyses (EFA) to identify latent constructs (principal axis factoring, Equamax rotation). Factor scores were included as sociobehavioral predictors in subsequent models. Spearman correlation was conducted to evaluate collinearity among predictors.

#### 2.5.3. Associations between predictors and cognitive outcomes

Linear mixed-effects models (LMM) were performed to identify factors influencing baseline or longitudinal (interaction term with time) cognition. Because we were mostly concerned with intra-individual changes over time, use of PC scores in LMM also reduces influence from one or more cognitive tests having greater fluctuation. Predictors were entered into the models as fixed factors or covariates. Time in months since the baseline visit was entered both as fixed and random effects, and subject’s random intercept was included to account for within-person clustering. A first-order autoregressive covariance structure was used to model the random effects. Models were built stepwise, such that all predictors were included in the initial model but were iteratively removed for p>0.15. The Akaike information criterion (ΔAIC>2) was used to decide if a model iteration was better than the previous with a preference for simpler model for ΔAIC≤2.

## 3. Results

PINE participants had relatively low socioeconomic status (SES), with average education of 9 years and 83.73% making less than $10,000 per year (Table 1). Most immigrated to the U.S. during middle age after 1980, and have been U.S. residents for almost two decades.

**Table 1.**
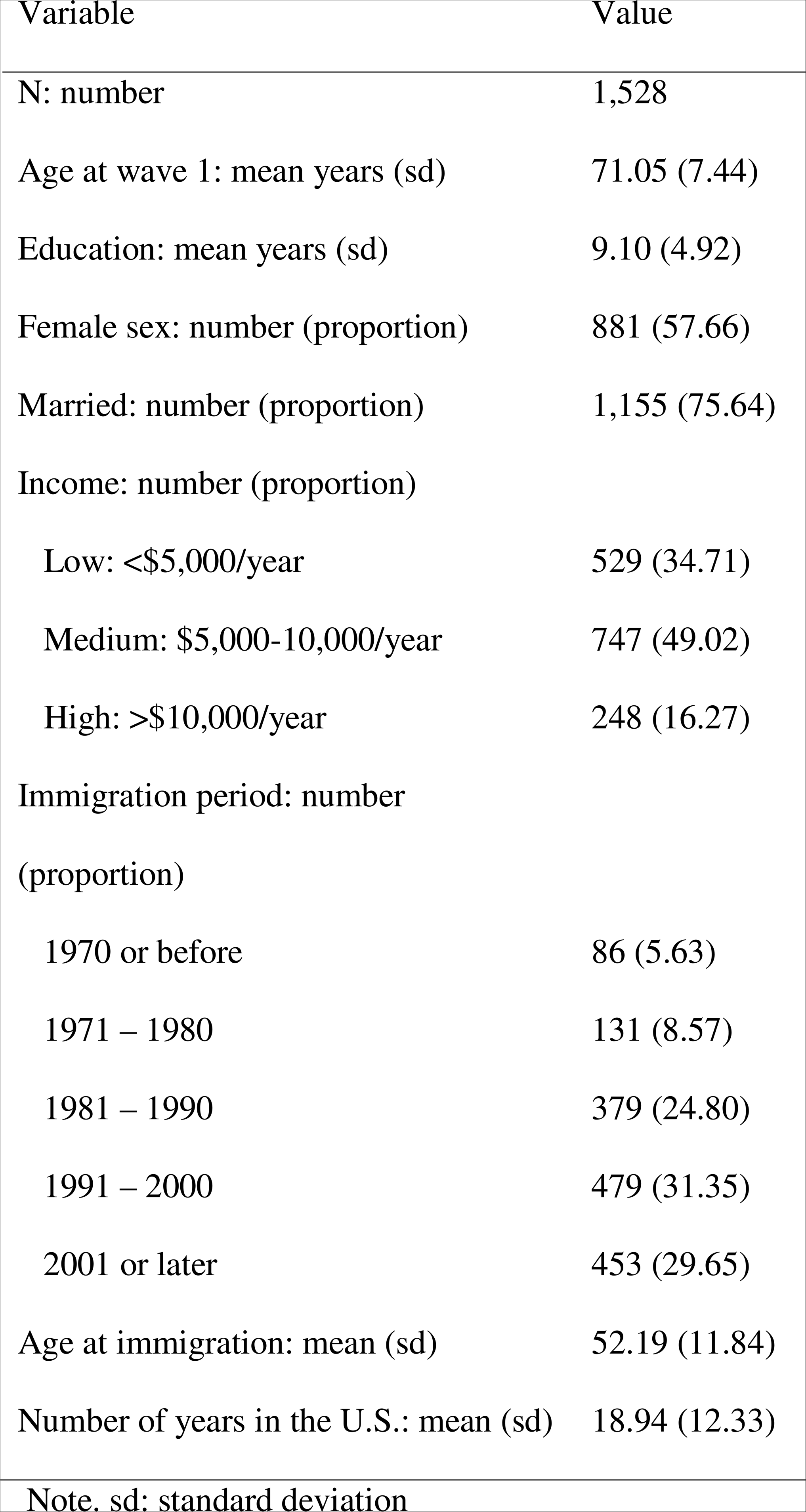
Demographic characteristics of the sample.

### 3.1. PCA of cognitive outcomes

In keeping with the need for rigorous analysis of cognition including deriving cognitive PC scores for analysis using LMM, up to 20% of participants’ cognitive scores fluctuated by one standard deviation or more between time points (Supplementary Figure 1). PCA of the five adjusted cognitive Z-scores showed Kaiser-Meyer-Olkin (KMO) sampling adequacy of 0.396 and Barlett’s test of sphericity at p<0.001. These modest correlations among the cognitive measures thus supported our selection of PCA over factor analysis for cognitive outcomes. Two PCs were extracted (eigenvalues>1.0, Figure 2 Panel B): EBMT-immediate recall, EBMT-delayed recall, and EBMT-percent retention loaded onto the memory PC; digit span backward and SDMT loaded onto the executive functioning PC.

**Figure 2.**
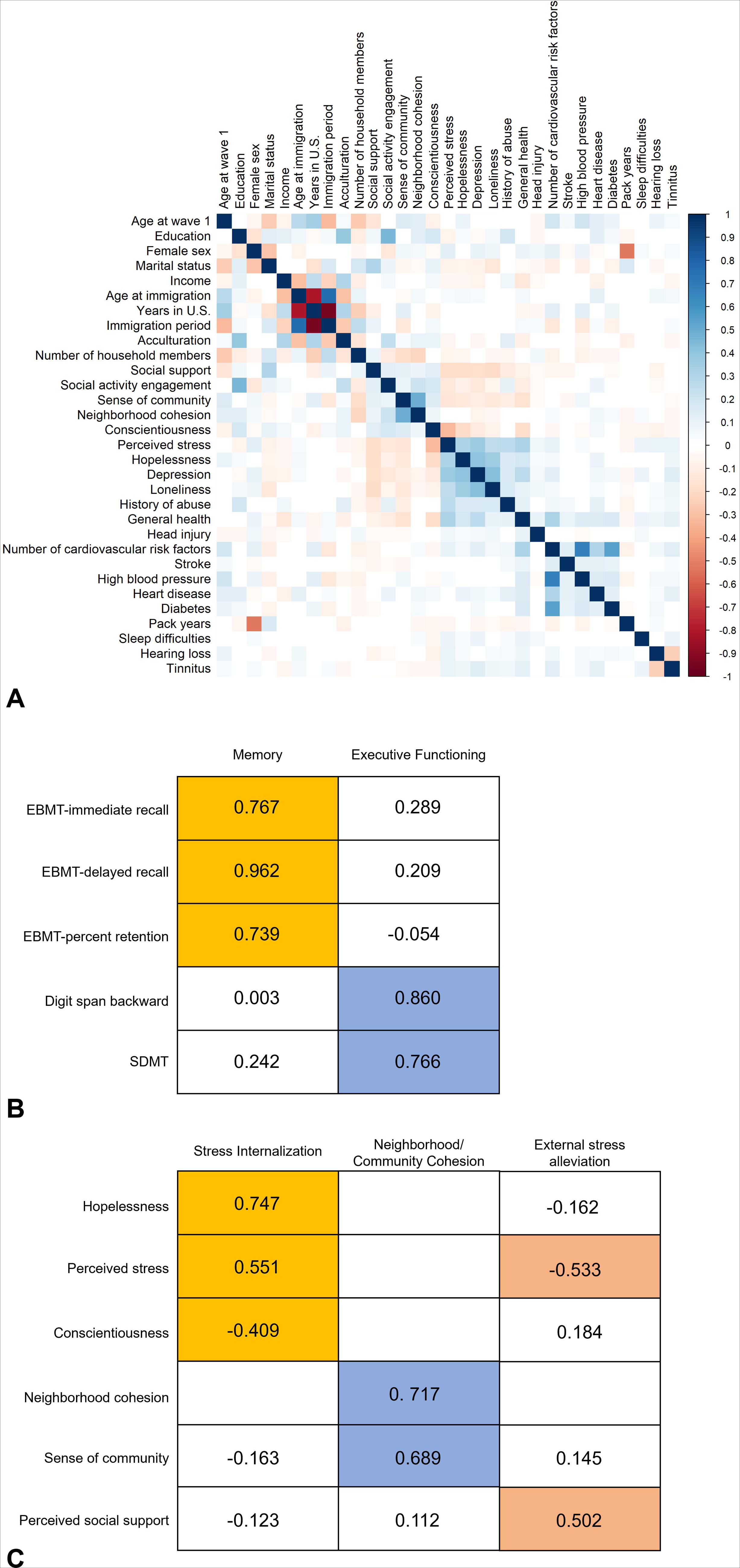
Correlations among predictors, and loadings for principal component and factor analyses. Note. Panel A illustrates the correlations among predictors. Panel B illustrates the strength of the component loadings for the cognitive variables from the rotated component matrix. Panel C illustrates the strength of the factor loadings for the psychosocial variables from the rotated factor matrix. EBMT: East Boston Memory Test; SDMT: Symbol Digit Modalities Test. P values are adjusted using the Benjamini-Hochberg correction, with the false discovery rate set at 0.05.^62^

### 3.2. EFA of psychosocial constructs

As predicted, bi-variate analysis showed multiple correlations among sociobehavioral variables (Figure 2 Panel A). EFA identified three factors (scree plot, two with eigenvalues >1.0; KMO of 0.624, Barlett’s test of sphericity p<0.001; Figure 2 Panel C). One factor (stress internalization) loaded onto measures of hopelessness, perceived stress, and lack of conscientiousness; a second factor (neighborhood/community cohesion) loaded onto neighborhood cohesion and sense of community. The third factor (external stress alleviation) loaded onto higher perceived social support and lower levels of perceived stress. No factor loaded onto social engagement and acculturation, and they were subsequently analyzed as independent variables.

### 3.3. Identifying significant predictors of cognitive outcomes

For memory, greater baseline PC score was associated with older age, fewer years of formal education, higher annual income, higher level of acculturation, and absence of strokes. Some of these differences dissipated over time, including effects from education and acculturation regressing towards the mean (Table 2, Figure 3, and Supplementary Table 2). Higher stress internalization and stroke were each associated with greater rates of memory decline over time. For each standard deviation increase in stress internalization, we observed a 0.024 unit greater annualized decline in memory function. History of stroke was associated with 0.084 unit greater annualized decline in memory, although the overall frequency of stroke diagnosis (n=50) was low so interpretation may be limited.

**Figure 3.**
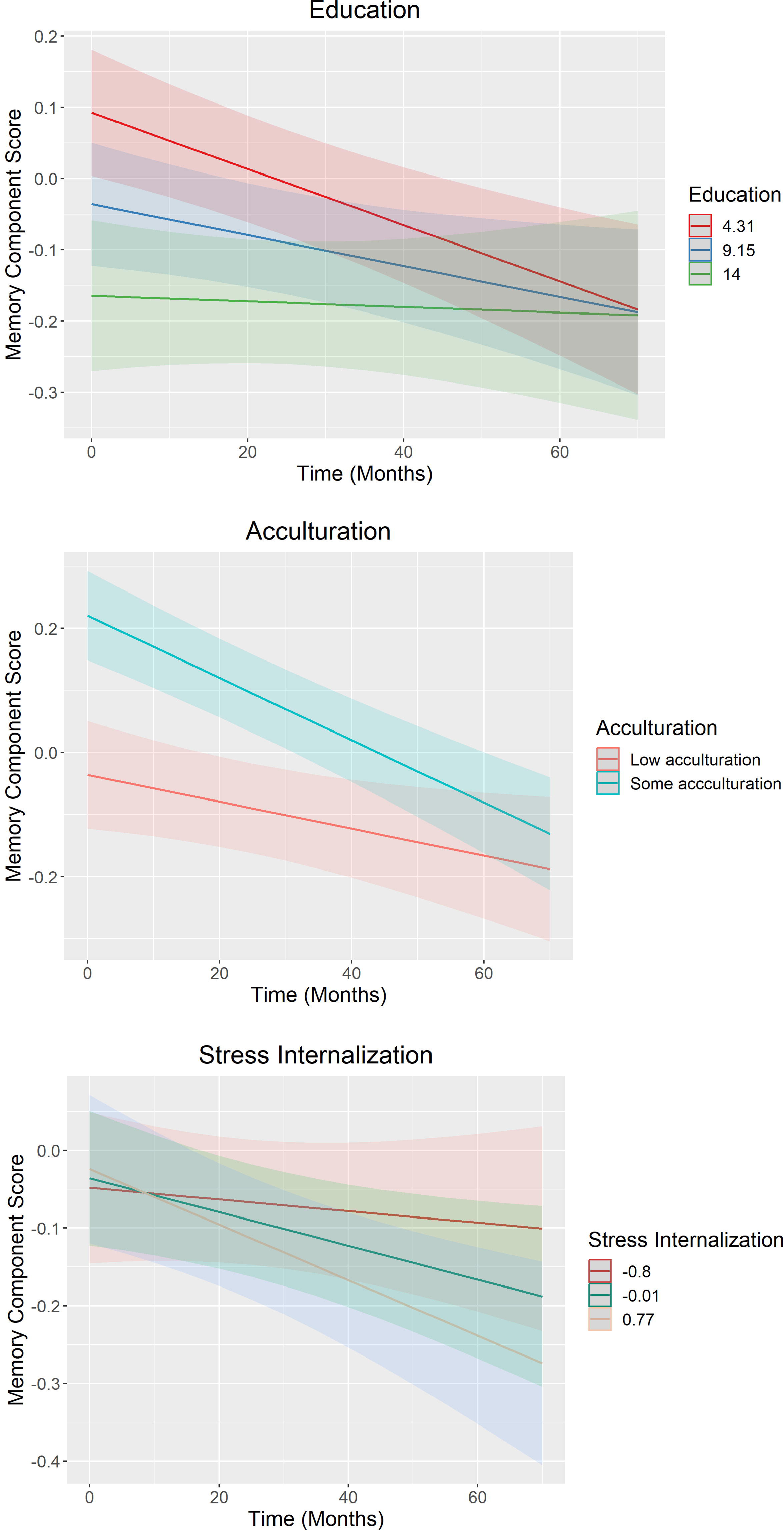
Significant longitudinal predictors of memory component scores. Note. Moderating effects of longitudinal predictors on decline on memory component score over time. Continuous moderators (education and stress internationalization) are stratified at the mean as well as one standard deviation above and below the mean.

**Table 2.**
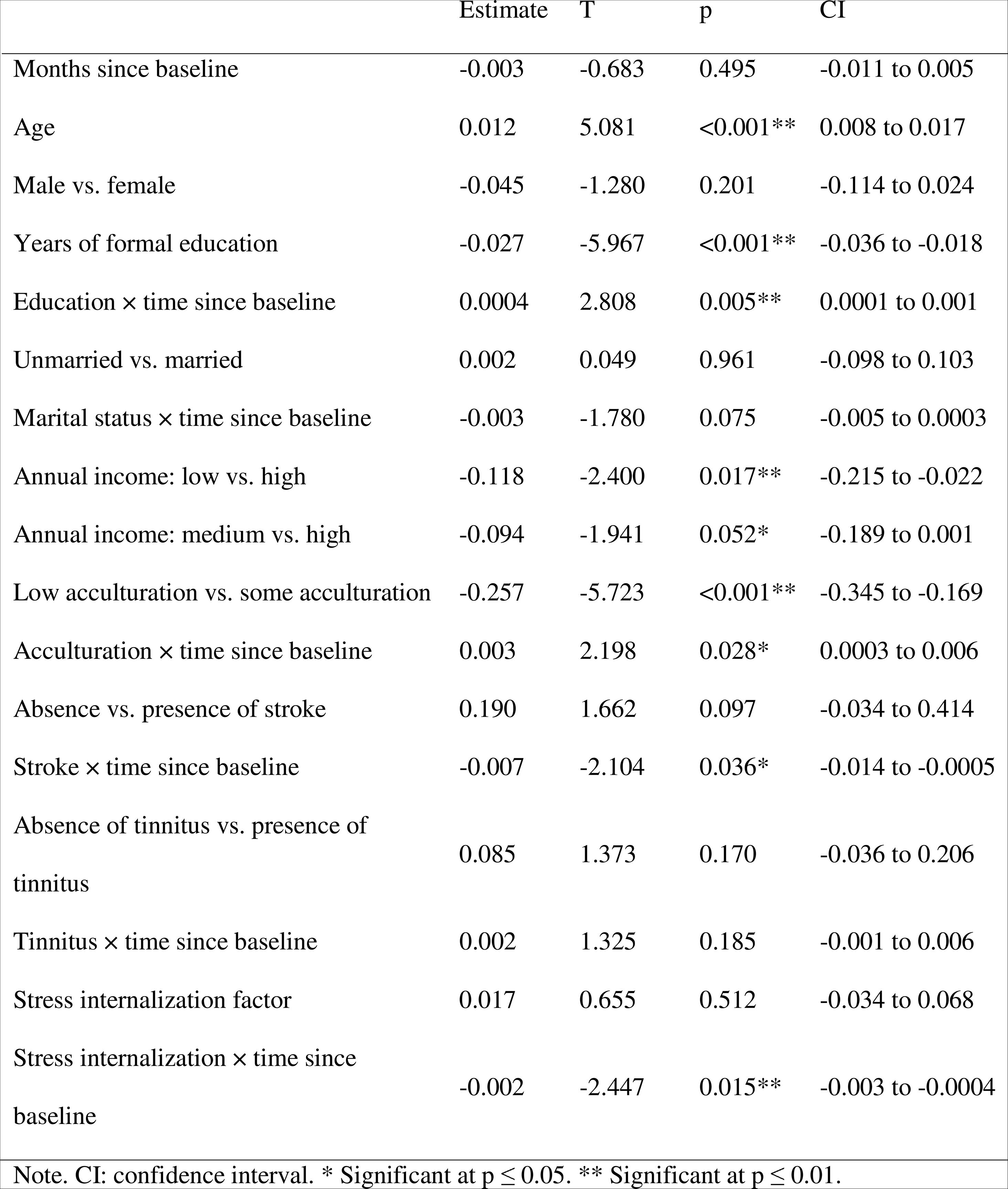
Estimates of fixed effects for the memory component.

We next examined factors associated with change in executive functioning over time. Multiple factors affecting baseline memory PC score also influenced executive functioning PC score, including age, formal education, annual income, and acculturation. Depressive symptoms, stroke, heart disease, and low social engagement but greater neighborhood/community cohesion factor were additionally associated with worse baseline executive function (Supplementary Tables 3 & 4). None of the clinical or sociobehavioral factors examined influenced longitudinal executive functioning, and only greater formal education was associated with greater decline in executive function (Supplementary Table 4). However, there was no difference in slope of unadjusted executive function scores over time among PINE participants (Supplementary Figure 2), and we suspect the effect of education on executive functioning to result from education-adjustment during Z-transformation. As this relationship was not observed for memory, we made the conservative conclusion that greater education in older U.S. Chinese was not protective against longitudinal decline in executive functioning.

## 4. Discussion

Among sociobehavioral factors examined in PINE, we identified three intrinsic constructs linking multiple survey outcomes: stress internalization, neighborhood/community cohesion, and external stress alleviation. Even when compared alongside demographic and clinical risk factors, stress internalization showed strong association with memory decline over three consecutive waves in PINE. In contrast, none influenced change in executive functioning over time.

Most studies leveraging the HAAS and the MESA have focused on biological predictors of cognitive decline. Participants in these studies either represent one of the most populous minority ethnic groups in the region (HAAS) or had high SES (MESA), and generalizability of these studies to U.S. mainland Asian Americans or Asians with lower SES is unknown. To our knowledge, only one previous PINE study examined cognitive trajectories in more than two waves.^18^ It identified a strong relationship between baseline function and rates of decline (e.g., those with worst baseline function had fastest cognitive decline). As we discussed, the group with worst baseline function and fastest temporal decline likely included participants with undiagnosed cognitive impairment. Sociobehavioral factors associated with their cognitive change may have more to do with behaviors during early stages of dementia than risk-associated behaviors predisposing to dementia onset. A corollary of this is the inclusion of participants with incident cognitive impairment among those with better baseline cognition. In keeping with these concerns, cross-sectional associations with cognition identified in prior PINE studies largely failed to influence longitudinal cognition in our analysis – due to fallacy of reverse causality, regression towards the mean, or both. Our study thus underscores the importance of leveraging longitudinal cognitive and functional assessment to retrospectively identify a baseline group with normal cognition,

The identification of multiple latent sociobehavioral constructs has conceptual and practical implications. It is not known if there exists more prominent clustering of sociobehavioral factors in older immigrants or minority adults than in older White adults. The first two groups’ neighborhood-level factors were best known to be more influenced by race relations (e.g., redlining) than choice.^41^ Behavioral clustering was also identified in geographically dispersed research participants according to income and cultural factors.^42^ The novel construct consisting of hopelessness, perceived stress, and low conscientiousness in PINE deserves follow-up investigation for its association with longitudinal memory decline. While we conceptualized this factor as stress internalization, these traits could also represent a culturally specific phenotype for depression or another mood disorder. In keeping with this, older Asian immigrants in North America – especially among those with lower levels of acculturation and English proficiency – face increased depression risks.^43^ Higher perceived stress itself is linked to lower baseline cognitive scores, faster cognitive, increased risk of dementia, and reduced brain volume.^44–47^ Hopelessness^48,49^ and low conscientiousness have also been respectively linked to cognitive impairment in the Finland-based Cardiovascular Risk Factors, Aging, and Dementia Study in Finland^48^ and the U.S.-based Religious Order Study.^50^ Whether these traits herald more severe depression^44^ or pre-symptomatic neurodegeneration^50^ remains to be determined, yet their replication across three very distinct cohorts deserves further mechanistic exploration. Because these traits correspond more to stress perception and processing than mere presence of stressors, their potentially modifiable nature can be a unique focus of future sociobehavioral intervention.

One potentially unique aspect of stress internalization among older Chinese adults involves the model minority stereotype. Asian Americans are often monolithically seen as being spared by health disparities,^51^ and older U.S. Chinese may thus feel the need to silently endure various psychosocial stressors such as lower English proficiency and acculturation, reduced participation in mainstream American society and healthcare, and discrimination. In the context of low SES, high-effort coping have been associated with worse health outcomes – a phenomenon known as John Henryism, originally posited to explain the high prevalence of hypertension among low resourced Southern Black men.^52^ While John Henryism has been primarily studied in Black Americans,^53^ the notion of high effort coping beyond available resources may similarly explain a maladaptive response to the model minority stereotype among PINE participants, especially if they regularly deal with greater stress than HAAS participants and with fewer resources than Chinese MESA participants. If this relationship is validated, efforts to alleviate the model minority stress may be more fruitful in preventing longitudinal cognitive decline in older U.S. Chinese than nebulous public health efforts focused on the built environment or community dynamics.

Notably, we failed to replicate in our longitudinal analysis many findings from cross-sectional analyses of the same cohort. This is not unexpected, as some factors (e.g., education, acculturation) may improve performance on baseline neuropsychological testing^54,55^ without altering longitudinal rates of change. Parallel to our observation, this phenomenon has been observed in U.S. Latino immigrants.^56,57^ However, limited benefits of other sociobehavioral factors may deserve a closer examination. For example, the role of social engagement on cognition is also more controversial in older Black^58,59^ and Japanese^60^ than White Americans.^61^ It remains possible that existing scales to assess social engagement in mainstream U.S. society do not fully capture the spectrum of leisurely or engaging activities among Hispanic, Asian, and Black Americans. For U.S. Chinese, affirmative responses to social engagement questionnaires may be limited by feasibility and affordability of overnight trips, eating out, or attending a concert for immigrants who do not drive. Lower English proficiency and personal preferences may further distance survey responses from social fulfillment or cognitive enrichment. Until more culturally-relevant scales for social engagement or support can confirm their protective effects against cognitive decline in U.S. Chinese, these factors should be reserved more for promotion of mental well-being than evidence-based prevention of cognitive aging.

Results from this study should be interpreted with the following limitations. While this was the largest community-based study of older U.S. Chinese, participants were all recruited from the Chicago metropolitan area and nearly 1000 individuals were lost to follow-up between waves 1 and 3. Sampling bias may thus limit our findings’ generalizability, but the consistency of our positive finding on stress internalization and memory with other large non-Asian cohorts was reassuring. Typical for community-based studies, our small cognitive battery may lack sufficient sensitivity for subtle cognitive decline. The brain-behavior relationships for memory and executive function components also have not been directly validated in a Chinese-speaking population, although we were successful in deriving two distinct cognitive PCs in keeping with their English originals. While many study instruments were commonly used in China-based cohorts and others and were forward/backward translated with the input of a CAB, their cultural appropriateness may lag behind linguistic accuracy. Many sociobehavioral variables were based on self-reports which can be subject to response or recall bias. Similarly, a diagnosis-based derivation of medical co-morbidities risks reverse causality related to health care access. Nevertheless, we present a novel yet robust finding in line with other large population-based non-Asian cohorts which should broadens its generalizability and should further prioritize it in future prevention studies.

## Funding

This study was funded by the Rutgers University Asian Resource Center for Minority Aging Research (RCMAR; P30 AG059304), Rutgers University Resource Center for Alzheimer’s and Dementia Research in Asian and Pacific Americans (RCASIA; P30 AG083257), and the Alzheimer’s Disease Neuroimaging Initiative (ADNI; U19 AG024904) Health Equity Scholar Program, which were all funded by the National Institute on Aging (NIA).

## Declaration of interest

WTH has patents on CSF-based diagnosis of FTLD-TDP and prognosis of MCI-AD and SMA; licensed COVID-19 serology tests to SigmaMillipore; and consulted for Apellis, Biogen, and Fujirebio.

## Data Availability

All data produced are available online at https://www.chinesehealthyaging.org/pinestudy.html

## Notes

### Author Declarations

IRBs of Rush University and Rutgers University gave ethical approval for this work.

