## Supplementary Materials for "Stress internalization associated with cognitive decline among older U.S. Chinese"

### Supplementary Table 1. Study Measures.

| **Variable** | **Original Measure** | **Response type** | **Notes on data transformations and modeling considerations** |
| --- | --- | --- | --- |
| Global cognition | Chinese version of the Mini-Mental State Examination (C-MMSE)^1^ | Continuous raw score | Used to determine eligibility criteria (C-MMSE>28 at wave 3) but not used for principal component analysis because this was a general cognitive measure and did not tap into specific cognitive domains. |
| Episodic memory | East Boston Memory Test (EBMT)^2^ | Age-, sex-, and education-adjusted Z scores | Episodic memory measure that included immediate and delayed recall components; a percent retention score was also calculated as the ratio of delayed recall to immediate recall×100 (capped at 125%). |
| Working memory | Wechsler Memory Scale-Revised (WMS-R)^3^ Digit Span Backward subtest | Age-, sex-, and education-adjusted Z scores |  |
| Processing speed | Symbol Digit Modalities Test (SDMT)^4^ | Age-, sex-, and education-adjusted Z scores |  |
| Instrumental activities of daily living (IADL) |  | Continuous | Lawton-Brody IADL^5^ score was calculated for each participant based on responses to corresponding PINE questions on whether the participant needed help with managing money, making a telephone call, preparing meals, doing laundry, taking medications, performing regular housework, shopping, traveling/commuting. This showed no difference in IADL between men and women, suggesting that the differential threshold approach previously attributed to Western gender-roles did not apply to this cohort. We thus used a baseline score of 7 or more (with 6.5 equivalent to Z-score of -1.5) as an indicator of normal function. |
| Age |  | Continuous in years |  |
| Education |  | Continuous in years |  |
| Marital status |  | Married or not married |  |
| Income |  | Low, medium, high | The majority of the sample reported lower income ranges, so reported income was divided into tertiles (<$5,000, $5,000-$10,000, and >$10,000). |
| Acculturation | PINE Study Acculturation Scale^6^ adapted from a short acculturation scale developed for Hispanics^7^ | Low acculturation, some acculturation | The sample largely reported low acculturation, so a median split was performed to dichotomize the variable into low acculturation (≤13; preferring mostly Chinese language, media, and friends; 36%) and some acculturation (>13; 64%). There was a survivorship bias such that those who immigrated earlier were older than those who immigrated more recently at study entry. Moreover, due to recruitment strategies focused on more recent immigrants, there was a significant association between age at baseline and age at immigration. Therefore, age at immigration was not included as an independent predictor. |
| Medical health conditions |  | Presence or absence | Participants were asked if they had been diagnosed by a medical professional for heart disease, stroke, cancer, high cholesterol, diabetes, high blood pressure, hip fracture, thyroid disease, and osteoporosis. |
| Number of cardiovascular risk factors |  | Continuous, 0-4 | Count of existing cardiovascular risk conditions, including heart disease, stroke, high blood pressure, and diabetes. |
| Review of systems (ROS) |  | Presence or absence | Participants were asked whether they were currently or had been treated for symptoms in 15 organ systems, including general, skin, head, ears, eyes, nose, throat, breasts, respiratory, cardiovascular, gastrointestinal, urinary, vascular, musculoskeletal, and neurologic. |
| Pack years |  | Continuous | Number of packs of cigarettes smoked per year multiplied by number of years smoked was calculated from participant’s report of cigarette use. |
| Depression | 8-item Patient Health Questionnaire (PHQ-8)^8^ | Low, medium, high | 8-item version of the depression inventory; scores were categorized into low, medium, and high based on established clinical ranges (0-4 for none-minimal, 5-9 for mild, 10-14 for moderate, 15-24 for severe)^8^ due to the vast majority of participants endorsing 0 which resulted in a highly skewed distribution. |
| Anxiety | Hospital Anxiety and Depression Scale (HADS)^9^ | Continuous | Anxiety items from the HADS were used in PINE; anxiety score was not ultimately included in the model due to low endorsement. |
| Neighborhood cohesion |  | Log- and Z-transformed | Interactions among neighbors were assessed using questions adapted from the Chicago Health and Aging Project.^10^ |
| Sense of community | Sense of Community Index^11^ | Z-transformed | 12-item survey measuring perception of belonging, influence, commitment, and connection to one’s community. |
| Conscientiousness | 12 items from the NEO Five-Factor Inventory^12^ | Z-transformed |  |
| Neuroticism | 6 items from the NEO Five-Factor Inventory^12^ | Z-transformed | Neuroticism was not ultimately included in the models because it was only a subset of the original scale of neuroticism and had shown reduced internal reliability in the PINE sample.^13^ |
| Perceived stress | Perceived Stress Scale (PSS-10)^14^ | Z-transformed | 10-item inventory measuring unpredictability, uncontrollability, and overloading of stress experienced. |
| Feelings of hopelessness | Beck Hopelessness Scale^15^ | Z-transformed | 7-item short form, which was developed and validated among patients with terminal cancer,^16^ measuring pessimistic cognitions. |
| Perceived social support |  | Z-transformed | Levels of perceived social support from family and friends were evaluated by questions from the Health and Retirement Study (HRS)^17^ |
| Social activity engagement |  | Z-transformed | 16 questions regarding frequency of engagement in various social activities (e.g., reading, playing mahjong, visiting relatives, friends, or neighbors), measured on a 5-point Likert scale.^18^ |

### Supplementary Table 2. Type III tests of fixed effects for the memory component.

|  | Numerator df | Denominator df | F | p |
| --- | --- | --- | --- | --- |
| Time since baseline | 1 | 1576.418 | 5.203 | 0.023* |
| Age | 1 | 1430.120 | 25.816 | <0.001** |
| Sex | 1 | 1408.864 | 1.638 | 0.201 |
| Years of formal education | 1 | 1844.794 | 35.600 | <0.001** |
| Education × time since baseline | 1 | 1624.848 | 7.884 | 0.005** |
| Marital status | 1 | 1863.511 | 0.002 | 0.961 |
| Marital status × time since baseline | 1 | 1594.939 | 3.169 | 0.075 |
| Annual income | 2 | 1405.605 | 2.940 | 0.053* |
| Stroke | 1 | 1804.840 | 2.763 | 0.097 |
| Stroke × time since baseline | 1 | 1562.901 | 4.426 | 0.036* |
| Tinnitus | 1 | 1812.922 | 1.886 | 0.170 |
| Tinnitus * × time since baseline | 1 | 1606.894 | 1.755 | 0.185 |
| Acculturation | 1 | 1808.387 | 32.749 | <0.001** |
| Acculturation × time since baseline | 1 | 1563.938 | 4.831 | 0.028* |
| Stress internalization factor | 1 | 1855.821 | 0.429 | 0.512 |
| Stress internalization × time since baseline | 1 | 1565.663 | 5.987 | 0.015** |

Note. * Significant at p ≤ 0.05. ** Significant at p ≤ 0.01.

### Supplementary Table 3. Type III tests of fixed effects for the executive functioning component.

|  | Numerator df | Denominator df | F | p |
| --- | --- | --- | --- | --- |
| Months since baseline | 1 | 1640.342 | 1.072 | 0.301 |
| Age | 1 | 1468.673 | 4.542 | 0.033* |
| Sex | 1 | 1455.341 | 1.600 | 0.206 |
| Years of formal education | 1 | 1837.768 | 6.272 | 0.012* |
| Education × months since baseline | 1 | 1730.497 | 14.790 | <0.001** |
| Marital status | 1 | 1454.411 | 3.122 | 0.077 |
| Annual income | 2 | 1446.844 | 5.139 | 0.006** |
| Acculturation | 1 | 1455.135 | 15.980 | <0.001** |
| Depressive symptoms | 2 | 3881.989 | 9.185 | <0.001** |
| Head injury | 1 | 1456.972 | 0.769 | 0.381 |
| Stroke | 1 | 1465.987 | 5.602 | 0.018** |
| CV profile | 4 | 1450.598 | 2.265 | 0.060 |
| Hypertension | 1 | 1453.307 | 2.367 | 0.124 |
| Heart disease | 1 | 1446.935 | 7.001 | 0.008** |
| Sleep difficulties from ROS | 1 | 1427.900 | 6.286 | 0.012** |
| Stress internalization factor | 1 | 1575.890 | 1.254 | 0.263 |
| Neighborhood/community cohesion factor | 1 | 1457.505 | 8.098 | 0.004** |
| Social engagement | 1 | 1455.464 | 51.693 | <0.001** |

Note. * Significant at p ≤ 0.05. ** Significant at p ≤ 0.01.

### Supplementary Table 4. Estimates of fixed effects for the executive functioning component.

|  | Estimate | T | p | CI |
| --- | --- | --- | --- | --- |
| Months since baseline | 0.001 | 1.036 | 0.301 | -0.001 to 0.003 |
| Age | 0.006 | 2.131 | 0.033* | 0.0005 to 0.012 |
| Male vs. female | -0.054 | -1.265 | 0.206 | -0.137 to 0.030 |
| Years of formal education | -0.014 | -2.504 | 0.012** | -0.024 to -0.003 |
| Education × months since baseline | -0.0004 | -3.846 | <0.001** | -0.001 to -0.0002 |
| Unmarried vs. married | -0.088 | -1.767 | 0.077 | -0.185 to 0.010 |
| Annual income: low vs. high | -0.170 | -2.929 | 0.003** | -0.284 to -0.056 |
| Annual income: medium vs. high | -0.170 | -2.989 | 0.003** | -0.282 to -0.059 |
| Low acculturation vs. some acculturation | -0.173 | -3.998 | <0.001** | -0.257 to -0.088 |
| Depressive symptoms: low vs. high | 0.188 | 4.286 | <0.001** | 0.102 to 0.273 |
| Depressive symptoms: medium vs. high | 0.145 | 3.286 | 0.001** | 0.058 to 0.231 |
| Absence vs. presence of head injury | 0.150 | 0.877 | 0.381 | -0.186 to 0.486 |
| Absence vs. presence of stroke | 0.261 | 2.367 | 0.018** | 0.045 to 0.476 |
| Number of cardiovascular risk factors: 0 vs. 4 | -0.006 | -0.055 | 0.956 | -0.235 to 0.222 |
| Number of cardiovascular risk factors: 1 vs. 4 | 0.125 | 1.190 | 0.234 | -0.081 to 0.331 |
| Number of cardiovascular risk factors: 2 vs. 4 | 0.090 | 0.897 | 0.370 | -0.107 to 0.287 |
| Number of cardiovascular risk factors: 3 vs. 4 | 0.186 | 1.801 | 0.072 | -0.017 to 0.388 |
| Absence vs. presence of hypertension | 0.084 | 1.539 | 0.124 | -0.023 to 0.191 |
| Absence or presence of heart disease | -0.156 | -2.646 | 0.008** | -0.272 to -0.04 |
| Absence vs. presence of sleep difficulties r | 0.556 | 2.507 | 0.012** | 0.121 to 0.992 |
| Stress internalization factor | 0.028 | 1.120 | 0.263 | -0.021 to 0.078 |
| Neighborhood/community cohesion factor | -0.071 | -2.846 | 0.004** | -0.12 to -0.022 |
| Social engagement | 0.172 | 7.190 | <0.001** | 0.125 to 0.219 |

Note. CI: confidence interval. * Significant at p ≤ 0.05. ** Significant at p ≤ 0.01.

### Supplementary Figure 1. Changes in cognitive test raw scores over time.

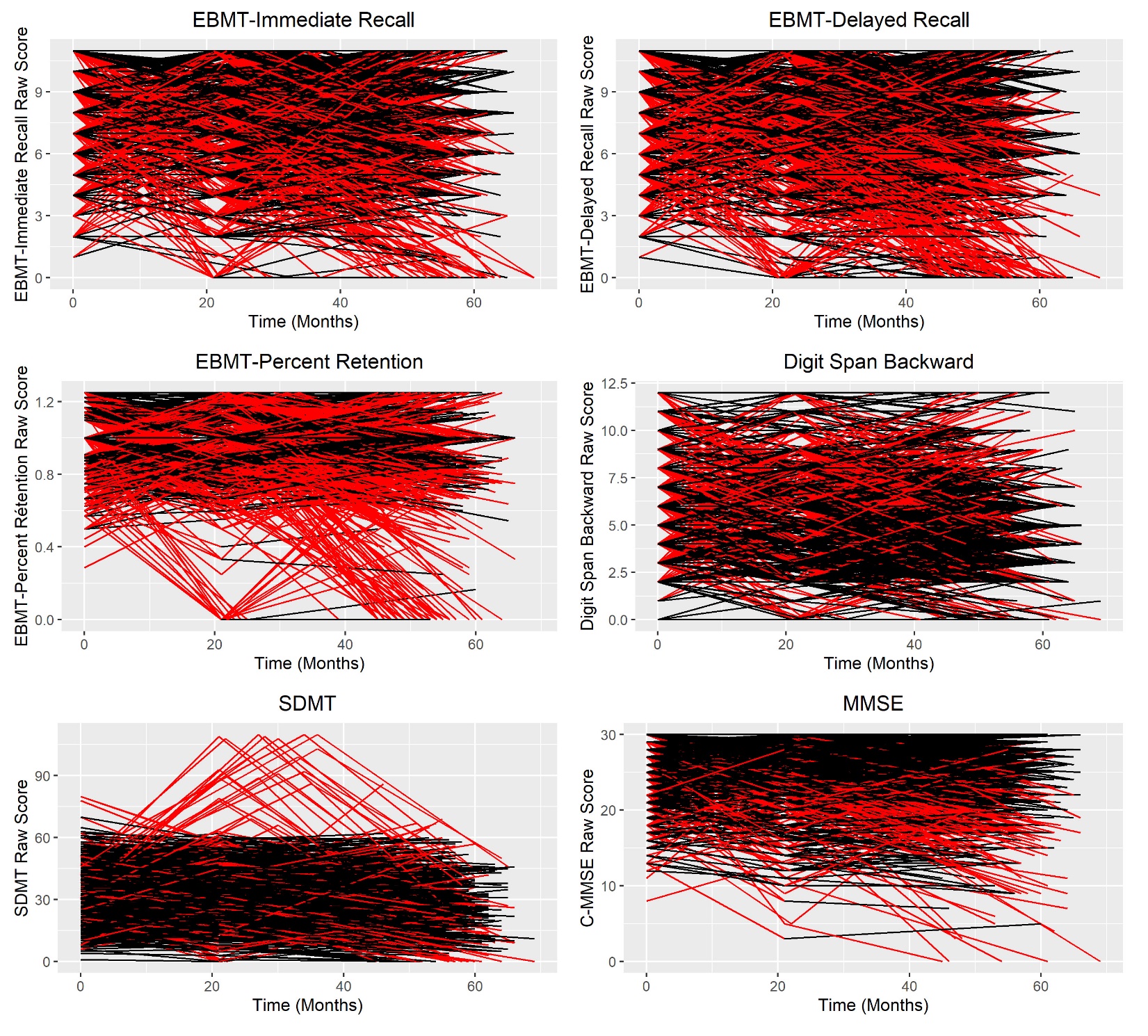

Note. Each line represents each participant’s cognitive trajectory across the three waves. Changes that were at least one standard deviation (either declined or improved) are highlighted in red. Significant changes constituted 8-20% of cases across cognitive measures. EBMT: East Boston Memory Test; SDMT: Symbol Digit Modalities Test; MMSE: Mini-Mental State Examination.

### Supplementary Figure 2. Moderating effects of education on changes in cognitive raw scores over time.

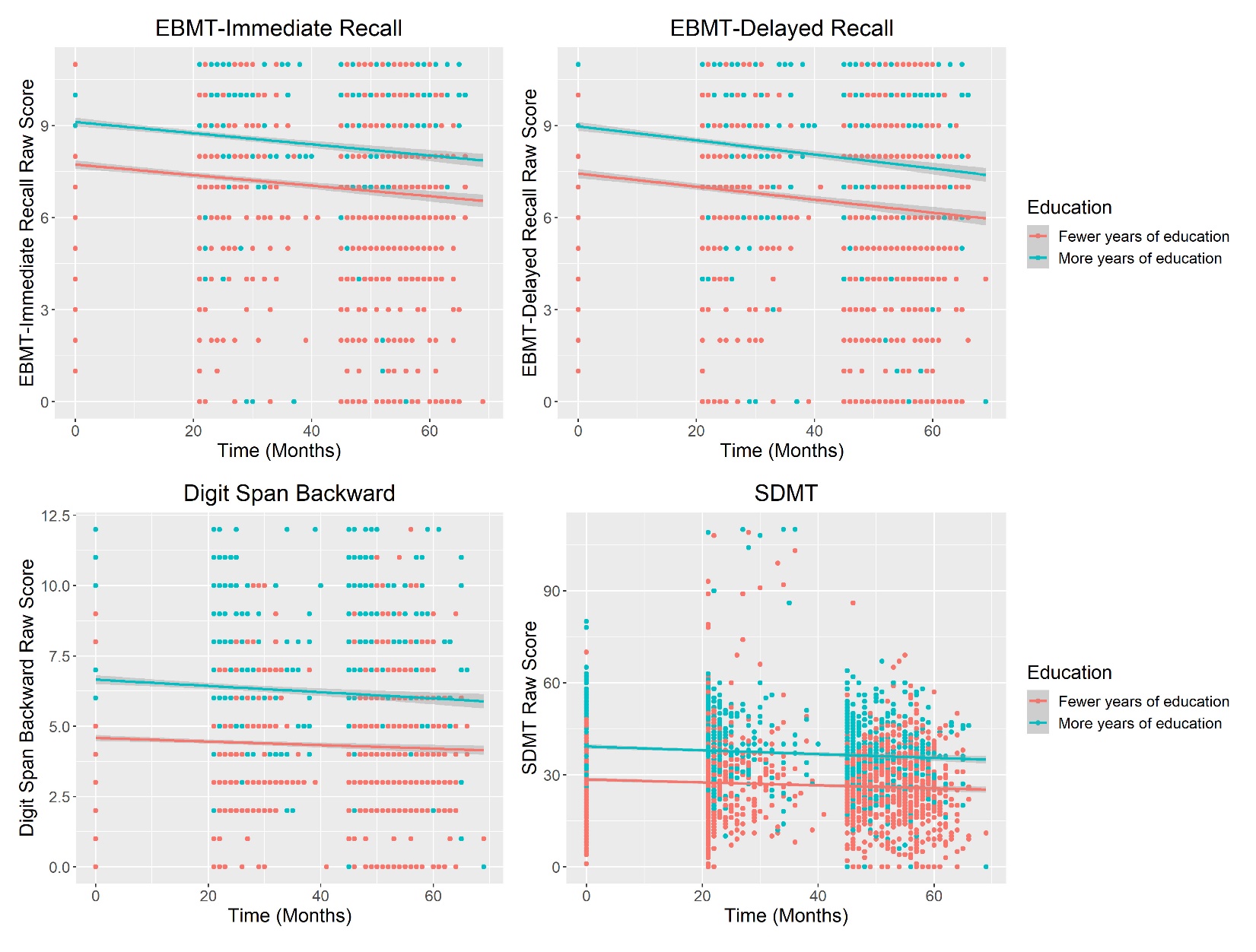

Note. Because the cognitive component scores were adjusted for education, we also examined the effects of education on the uncorrected cognitive raw scores, which showed generally better performance among those with more years of education, but the rates of decline were the same across educational groups. This suggests that significant interactions among memory and executive functioning component scores with time were likely an artifact due to education correction of the cognitive component scores, rather than true effects on longitudinal decline. EBMT: East Boston Memory Test; SDMT: Symbol Digit Modalities Test.
